# Exploring the correlation between COVID-19 fatalities and poor WASH (Water, Sanitation and Hygiene) services

**DOI:** 10.1101/2020.06.08.20125864

**Authors:** Godfred Amankwaa, Christian Fischer

## Abstract

Access to safe Water, Sanitation and Hygiene (WASH) services have been recognized as a highly precautionary measure essential to protecting human health during this COVID-19 outbreak. However, it is currently unknown *whether* poor or non-availability of these services are also closely related to COVID-19 fatalities. We analysed the latest data on COVID-19 fatality rates in Sub Saharan Africa with indicators of safe water and sanitation governance to test this hypothesis. We found a strong correlation between a higher case fatality rate and poorer access to safe drinking water as well as safe sanitation. The Pearson correlation is stronger for access to safe sanitation (−0.30) compared to access to safe drinking water (−0.20). The Chad, Niger and Sierra Leone were amongst the countries with the highest fatality rates (>6.0) and also had particularly poor access to safe drinking water (<34%) and safe sanitation (<22%). The hypothesis of an association between COVID-19 fatalities and poor access to *water and sanitation* was confirmed by this study. However, our analysis does not establish causality. Given the increase spread of COVID-19 and related deaths, this analysis serves as an important reminder that safe water and sanitation services are key for public health interventions and highlights the need to prioritise this sector in all economies.

## Introduction

The novel coronavirus (COVID-19) is spreading across countries and within countries. It is estimated that up to 70% of a population are likely to be infected with the coronavirus disease [1]. As of May 22, 2020, almost 334 173 people had died globally with the USA recording 96354 deaths [2]. While COVID-19 poses a massive crisis in the West, including deaths and the economic downturn, the nature of essential service provision in the global South presents some *informed fears* with regards to its spread and potential fatalities. For instance, ensuring good and consistent provision of safe water, sanitation, and hygienic services, such as regular water supply, availability of hand washing facilities, good WASH systems in health care facilities as well as good sanitation practices are essential to protecting human health during this COVID-19 outbreak [3]. Although the above highlighted measures are vital to keep up the fight against the spread of COVID-19, can the non-availability of these services be linked to potential COVID-19 deaths?

Considering the stark inequalities in water and sanitation provision, increased vulnerability and fragility of WASH infrastructure in most of the global South, we ask; *“Are people(households) with poor access to safe water and sanitation services more vulnerable to COVID-19 fatalities?”*. Currently, COVID-19 deaths have been associated with mortality conditions such as cardiovascular disease, diabetes, and hypertension etc and are more pronounced among the aged [4]. Yet, there is no empirical evidence on the relationship between COVID-19 deaths and access to water and sanitation.

Drawing on COVID-19 cases in Sub-Saharan Africa, we explored the hypothesis that; *lack or poor access to water and sanitation services are likely to increase potential COVID-19 deaths*. It is our hope that findings from this analysis will provide information to the global community about predicting the conditions of COVID-19 and access to safe water and sanitation. This will also feed into ongoing WASH interventions by most global South governments towards the fight against COVID-19 and other pandemics.

## Methods

To test this hypothesis, we obtained the latest data on COVID-19 fatalities from Our World in Data [5] and merged with some of the indicators present in the quality of governance dataset from Teorell et al. (2018) [6]. Among others, the Teorell et al’s [6] dataset contains the definition of ‘safe’ access to water and sanitation from the Environmental Performance Index of Hsu et al. [7]. Compared to the World Development Indicators’ notion of ‘*improved*’ access, ‘*safe*’ access is a more rigorous definition as water quality assessments also rest on the assumption that ‘improved’ water supplies are safe, but a significant amount of water supplies that meet the definition of an ‘improved’ source still do not meet WHO guidelines, e.g. water supplied through pipes may be contaminated, and groundwater may also be contaminated by faulty latrines, or the treatment of the water is inadequate. Sub-Saharan Africa was chosen to test this hypothesis because; (i)First, the region is the poorest including access to water and sanitation services [8]. It is estimated that of the 783 million people who are without access to clean water, 40% live in Sub-Saharan Africa and more than 320 million people lack access to safe drinking water [9] (ii) Secondly, an estimated 70% to 80% of the regions diseases are attributable to poor water quality [10] and (iii)lastly, the subsample of countries is relatively homogenous across many variables that potentially drive fatality rates, apart from access to safe sanitation and drinking. Univariate (Pearson correlation) analysis was conducted to test the hypothesis using R-studio.

## Results

Figure 1 and 2 summarises the relationship between case fatality rates and access to safe water and sanitation. A quick analysis illustrates a strong correlation between a higher case fatality rate and poorer access to safe drinking water as well as safe sanitation (see Figures below). South Africa has, as of May 20th, reported the highest fatalities (302), followed by Nigeria (192). The correlation is stronger for access to safe sanitation with a Pearson correlation test yielding -0.30 compared to - 0.20 for access to safe drinking water, which is intuitive. The Chad, Niger and Sierra Leone are amongst the countries with the highest fatality rates (>6.0) and also have particularly poor access to safe drinking water (<34%), safe sanitation (<22%). <u>Appendix A</u> and <u>B</u> provide an interactive version of the results in more detail.

**Figure 1:**
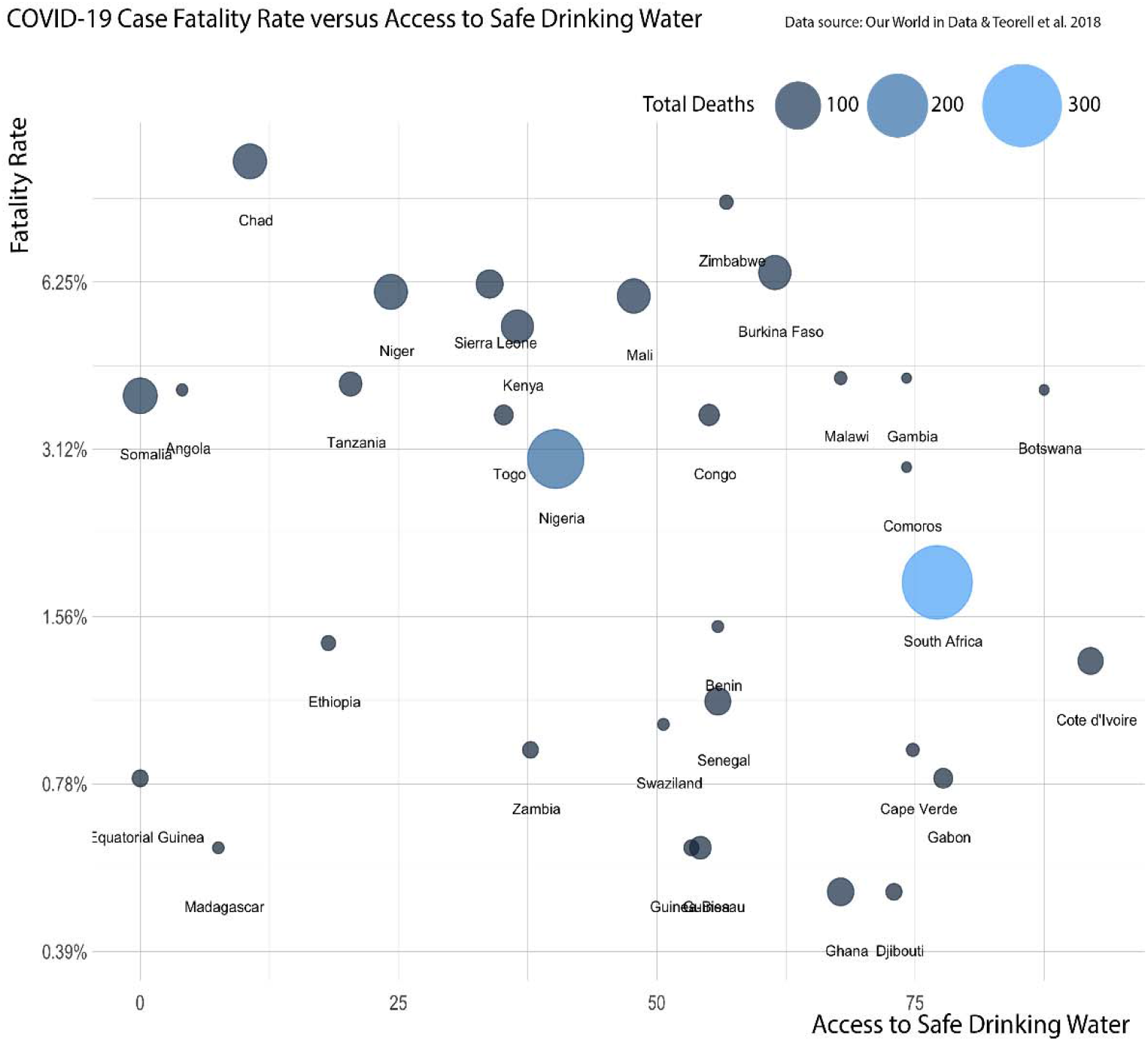
Correlation between COVID-19 case fatality rate and access to safe drinking water

**Figure 2:**
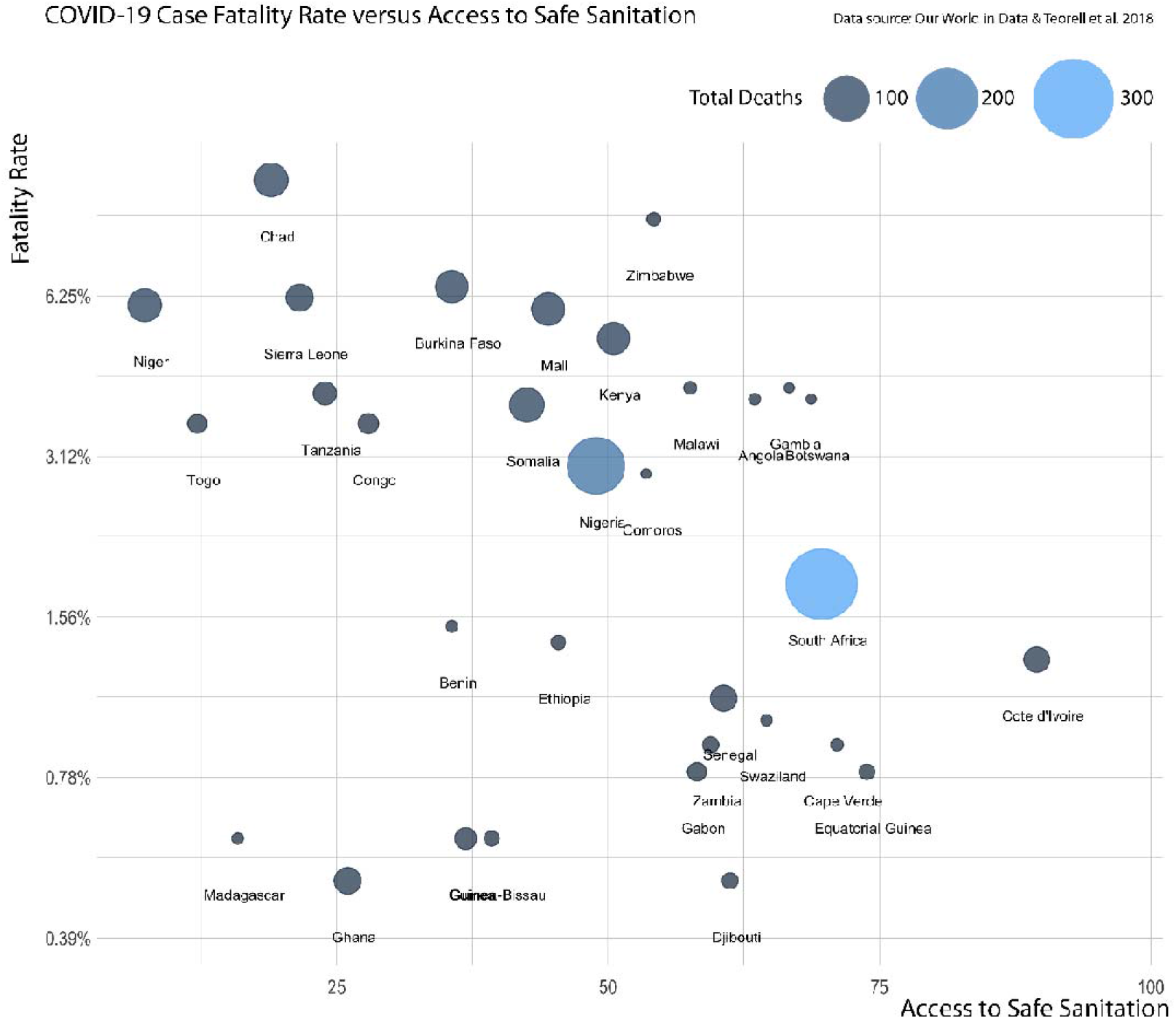
Correlation between COVID-19 case fatality rate and access to safe sanitation

## Discussion and conclusion

The hypothesis of an association between COVID-19 fatalities and poor *access to water and sanitation* is confirmed by the results. The above-mentioned analysis show that the case fatality rate is associated with the high percentage of poor access to safe sanitation and drinking water. This is consistent with the observation that handwashing with soap and water are protective measures against increased mortality rates during a crisis impacting most vulnerable populations. While other factors, such as median age, the quality of health services, the resiliency of an economy and above all, the scale of testing and reporting drives the number of fatalities, the sub-Saharan cluster of countries poses an arguably homogenous sample across many of these variables, with relatively young populations, and comparably poor health services and likely undertesting. The fact that we found such strong correlations between the total COVID-19 fatality rate and access to safe drinking water and safe sanitation is a strong indicator of the pivotal role that basic access to these services plays on an everyday basis. Most often too, WASH services are lacking even in the places where people go to seek treatment. With 57.5% of the Sub Saharan African population being multidimensionally poor and 492 million of them deprived of water [8] people are likely more exposed and vulnerable to the risks (*such as deaths*) of COVID-19 and other pandemics. The correlation is therefore plausible given that unsafe sanitation is responsible for 775,000 deaths each year and is the leading risk factor for infectious diseases [11]. This finding underscores the improvements that ought to be made in terms of equitable and safe services, only then can the overall disease burden be sustainably reduced, and the potential deaths associated with WASH be prevented.

We are not by anyway suggesting that our analysis is robust (*as it cannot establish causality between existing COVID-19 deaths and poor water and sanitation services*) nor insinuate that Sub-Saharan African countries may inevitably experience apocalypse as a result of the virus outbreak. However, we are using the trend in this brief analysis as a reminder of the importance of safe and improved WASH services not just for the fight against pandemics but in our daily lives. Apart from preventing potential deaths, many co-benefits will be realized by providing safely managed water and sanitation services and applying good hygiene practices. Times have changed and the current WASH responses for COVID-19 can be a turning point in alleviating the inequalities and poverty that characterise the water and sanitation sector. However, this demands bold policies and actions by all actors (governments, World agencies, utilities etc). COVID-19 is likely to hit the fragile economies in the global South harder and it is our hope that it forces countries to rapidly readjust their resiliency and emergency planning particularly for the water and sanitation sector. Although many national governments are shifting policies and keeping WASH priorities through the lens of COVID-19, adopting blended finance, digital water solutions and establishing low-income WASH response institutions will be useful for all [12]. Whilst our study focused on Sub-Saharan Africa, further studies can draw on additional data and other covariates to explore other key determinants more in-depth.

In conclusion, we demonstrate for the first time that poor access to safe drinking water and sanitation appeared to have an association with COVID-19 fatalities. Although our evidence does not establish causality, it specifically highlights the need for strong policies supporting WASH interventions, prioritising low and marginal populations in both developing and developed countries. It is critical for countries to understand that increasing safe water and sanitation services especially to the unreached will lead to better and healthier populations. Work presented here underscores the importance of all countries sector-specific response plans to prioritise WASH because a *WASH crisis is a health crisis*.

## Data Availability

All data used are included in the manuscript

## Supporting Data

<u>Appendix 1: Interactive version of COVID-19 vs access to safe drinking water</u>

<u>Appendix 2: Interactive version of COVID-19 fatality rate vs access to safe sanitation</u>

## Additional Data

Scholars who wish to use this dataset in their research are kindly requested to cite both the original source (as stated in this <u>Codebook</u>) and use the following citation: *Teorell, Jan, Stefan Dahlberg, Sören Holmberg, Bo Rothstein, Natalia Alvarado Pachon & Richard Svensson. 2018. The Quality of Government Standard Dataset, version Jan18. University of Gothenburg: The Quality of Government Institute, https://qog.pol.gu.se/doi:10.18157/QoGStdJan18*

## Declaration

We declare no competing interest

## Author Contributions

All authors contributed equally

## References

[1] CBS News. Coronavirus may infect up to 70% of world populations, experts warns. 2020 Available from https://www.cbsnews.com/news/coronavirus-infection-outbreak-worldwide-virus-expert-warning-today-2020-03-02/.

[2] Our World in Data, Statistics. 2020, Available from https://ourworldindata.org/coronavirus

[3] WHO. Water, sanitation, hygiene, and waste management for the COVID-19 virus. Interim guidance Report. 2020, April. Available from https://apps.who.int/iris/bitstream/handle/10665/331846/WHO-2019-nCoV-IPC_WASH-2020.3-eng.pdf?ua=1

[4] CDC. People Who Are at Higher Risk for Severe Illness. 2020, Available from https://www.cdc.gov/coronavirus/2019-ncov/need-extra-precautions/people-at-higher-zzrisk.html?CDC_AA_refVal=https%3A%2F%2Fwww.cdc.gov%2Fcoronavirus%2F2019-ncov%2Fhcp%2Funderlying-conditions.html

[5] Our World in Data, the Data. 2020. Available from https://ourworldindata.org/coronavirus-data

[6] Teorell J, Kumlin S, Dahlberg S, Holmberg S, Rothstein B, Alvarado Pachon N, Svensson R. The Quality of Government OECD Dataset, Version Jan18. University of Gothenburg: The Quality of Government Institute. 2018. Available from https://qog.pol.gu.se/

[7] Hsu, A., Etsy, D. C., Levy, M., de Sherbinin, A. Environmental Performance Index (Tech. Rep.). 2016. doi: doi:10.13140/RG.2.2.19868.90249

[8] Alkire, S., Dirksen, J., Nogales, R., Oldiges, C. Multidimensional poverty and COVID-19 risk factors: A rapid overview of interlinked deprivations across 5.7 billion people. 2020; OPHI Briefing 53, Oxford Poverty and Human Development Initiative, University of Oxford.

[9] UNICEF and WHO. Progress on household drinking water, sanitation and hygiene 2000-2017. Special focus on inequalities. New York: 2019, United Nations Children’s Fund (UNICEF) and World Health Organization, 2019.

[10] United Nations. Sustainable Development Goal 6 Synthesis Report 2018 on Water and Sanitation. New York, 2018. Available from https://www.unwater.org/publication_categories/sdg-6-synthesis-report-2018-on-water-and-sanitation/

[11] GBD 2017 Risk Factor Collaborators. Global, regional, and national comparative risk assessment of 84 behavioural, environmental and occupational, and metabolic risks or clusters of risks for 195 countries and territories, 1990–2017: a systematic analysis for the Global Burden of Disease Study 2017. Lancet (London, England). 2018 Nov 10;392(10159):1923.

[12] Amankwaa, G. COVID-19 and “Chasing for Water” - Water Access in Poor Urban Spaces. 2020 Available from https://iwa-network.org/covid-19-and-chasing-for-water-water-access-in-poor-urban-spaces/

